# Bourbon virus transmission, New York State

**DOI:** 10.1101/2022.10.28.22281663

**Authors:** Alan P. Dupuis, Melissa A. Prusinski, Collin O’Connor, Joseph G. Maffei, Cheri A. Koetzner, Tela E. Zembsch, Steven D. Zink, Alexis L. White, Michael P. Santoriello, Christopher L. Romano, Guang Xu, Fumiko Ribbe, Scott R. Campbell, Stephen M. Rich, P. Bryon Backenson, Laura D. Kramer, Alexander T. Ciota

**Author notes:** These authors contributed equally to this article (Alan P. Dupuis, Melissa A. Prusinski). **Address for correspondence:** Alan P. Dupuis II, The Arbovirus Laboratory, New York State Department of Health, 5668 State Farm Rd., Slingerlands, NY 12159.

## Abstract

In July 2019, Bourbon virus RNA was detected in an *Amblyomma americanum* tick removed from a resident of Long Island, New York. Tick infection and white-tailed deer (*Odocoileus virginianus*) serosurvey results demonstrate active transmission in New York, especially Suffolk County, emphasizing a need for surveillance anywhere *A. americanum* is reported.

## Introduction

Bourbon virus (BRBV, *Orthomyxoviridae*; *Thogotovirus*) is a suspected tick-borne human pathogen first isolated in 2014 from a patient that resided in Bourbon County, Kansas (1). BRBV is closely related to Oz virus, recently isolated from *Amblyomma testudinarium* ticks in Japan (2, 3). Since the initial discovery of BRBV, human cases have been identified in Kansas, Missouri, and Oklahoma (4). *Amblyomma americanum* (lone star tick) have been identified as the likely vector of BRBV transmission and maintenance (5, 6). Small and medium-sized mammals and ground dwelling birds such as wild turkeys (*Meleagris gallopavo*) are hosts for the immature ticks. Adults feed on large mammals, such as coyotes (*Canis latrans*) and white-tailed deer (*Odocoileus virginianus*). All three active developmental stages will bite humans (7). Virus detection in ticks and serologic evidence in mammalian hosts, including white-tailed deer, has been demonstrated in Missouri, Kansas, and North Carolina (6, 8-10).

## The Study

In July 2019, New York State Department of Health (NYSDOH) epidemiologists were notified that BRBV RNA was detected in an individual, partially engorged female *A. americanum* removed from a Long Island, NY resident. In addition to BRBV, the tick was positive for *Ehrlichia ewingii*. Comprehensive tick testing was performed at the University of Massachusetts (tickreport.com). Notes on the tick submission form indicated the resident was experiencing fever, chills, and fatigue. In response to this report, officials with NYSDOH and Suffolk County Department of Health Services (SCDHS) attempted to contact the resident for a follow-up investigation. No additional information was provided, and no blood samples were available to assess potential infection with BRBV.

In 2016, NYSDOH and SCDHS initiated active tick surveillance targeting *A. americanum* for BRBV and Heartland virus (HRTV). HRTV-infected ticks and seropositive deer were first detected on Long Island, NY, in 2018 and reported in 2021 (11). Standardized flag sampling was used for the collection of host-seeking *A. americanum* on public lands in Suffolk County. Between 2016-2020, 1,265 pools representing 4,189 adults, 7,227 nymphs, and 97 larvae tested negative for BRBV RNA by real-time reverse transcription PCR using an in-house developed multi-plex assay to detect HRTV and BRBV (11). The BRBV primers for this assay were designed based on the St. Louis strain, GenBank: Accession #MK453528 (12). During 2021, sampling for *A. americanum* on Long Island was expanded to collect a greater number of ticks from more locations and molecular detection protocols were modified to utilize BRBV-specific primers developed at TickReport (Amherst, MA) (Table 1). BRBV-specific primers were designed based on the original virus strain deposited in GenBank, Accession # KU708254 (13). A total of 1,058 pools, consisting of 4,406 adults (460 pools) and 9,972 nymphs (598 pools) were collected from 12 sites in Suffolk County. Pool sizes ranged from 1-10 adults and 5-20 nymphs. BRBV RNA was detected in five pools of unengorged nymphs. Positive pools were collected at a single site in Smithtown (*n*=3) on 3 May 2021 and two sites, one positive pool each, in Brookhaven on 9 June 2021 and 8 July 2021. Infectious virus was isolated from all BRBV RNA-positive tick pools after incubation on Vero cells (ATCC, Manassas, VA). The isolates were confirmed as BRBV by real-time reverse transcription PCR.

**Table 1.**
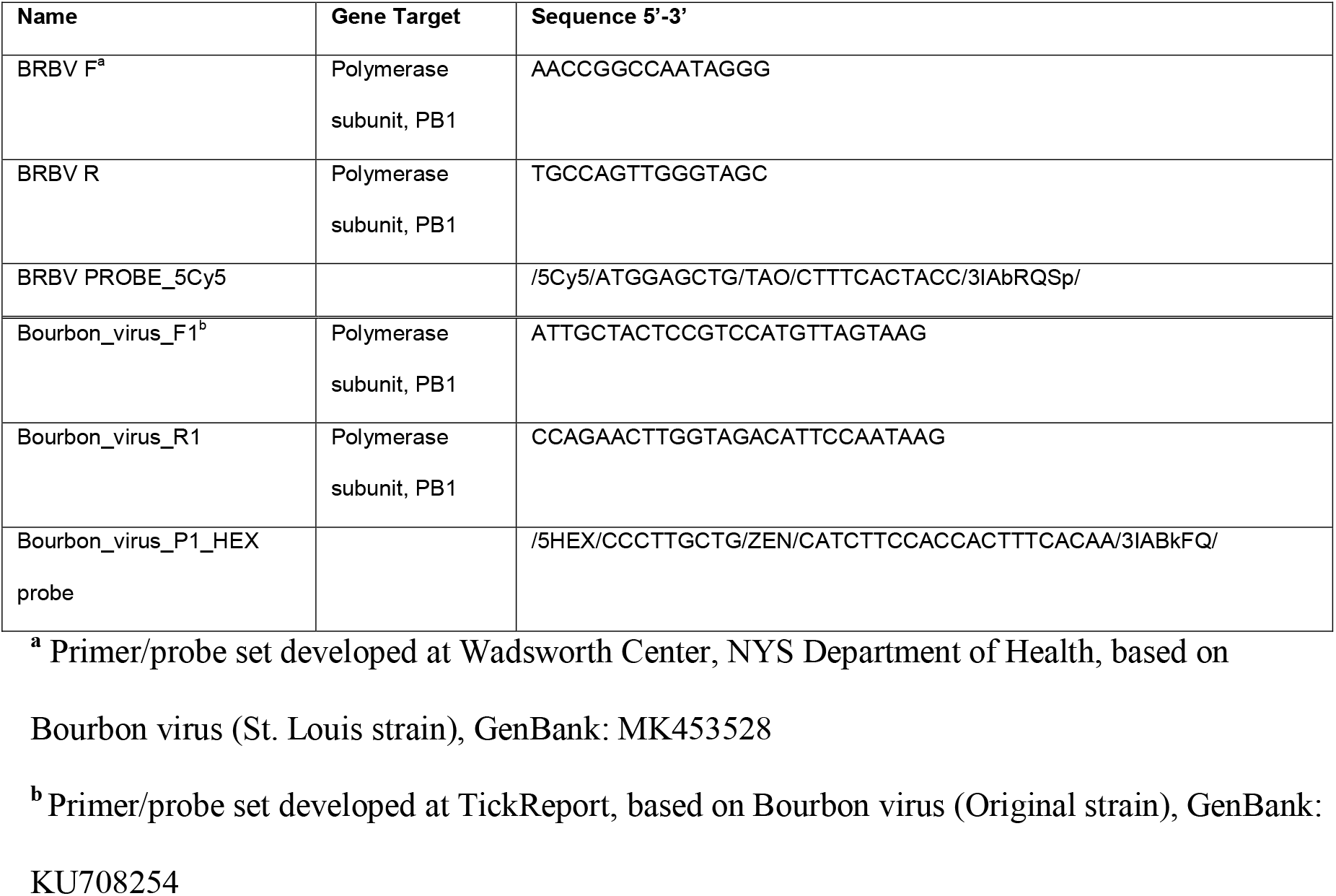
Primer/probe for detection of BRBV RNA

Serologic testing of hunter-harvested white-tailed deer blood submitted for arbovirus serosurveys has been conducted in NY since 2007 using the PRNT90, as described (14). BRBV was included in these assays starting in 2019 for deer harvested in Suffolk County and the Hudson Valley Region and 2020 for deer harvested in central and western NY. A total of 881 sera were screened at 1:20 for the presence of neutralizing antibodies to BRBV (Table 2, Figure 1). Positive sera were serially diluted for endpoint titers. Statewide, 37.7% of the deer were seropositive with titers ranging from 20 to >640, and 89.2% of the seropositive deer had titers greater than 20. The seropositivity was 66.5% for deer harvested in Suffolk County (Table 2, Figure 2). Seroprevalence was lower in western NY (3.8%), the Hudson Valley (1.7%), and central NY (1.2%). BRBV neutralizing antibodies were not detected in seven deer harvested in the northern NY region (Table 2, Figure 1). *A. americanum* ticks (*n*=1,641) collected from 145 deer harvested from Suffolk County were tested for BRBV RNA. The virus was not detected.

**Table 2.**
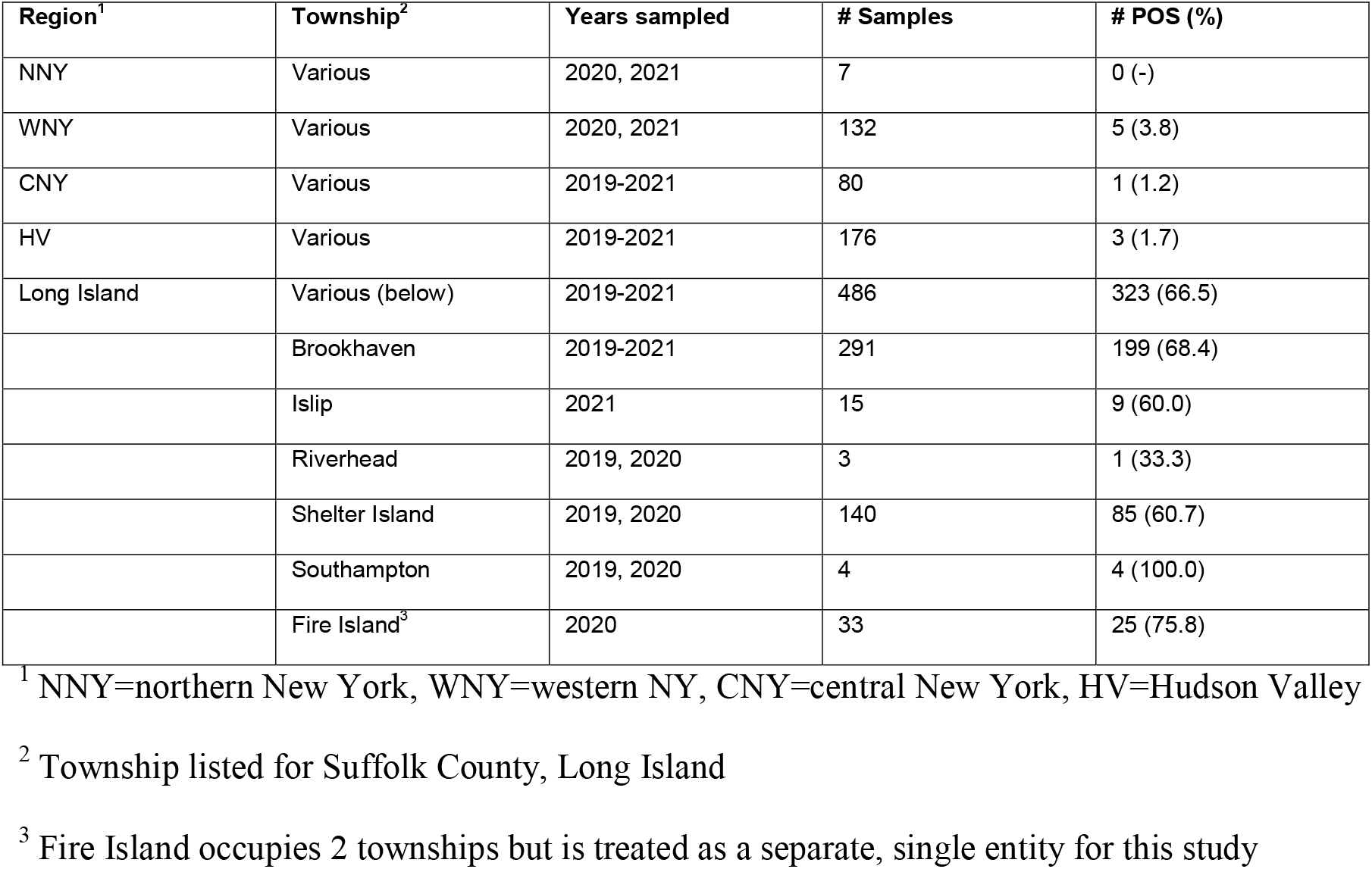
Plaque reduction neutralization test results for white-tailed sampled, New York State

**Figure 1.**
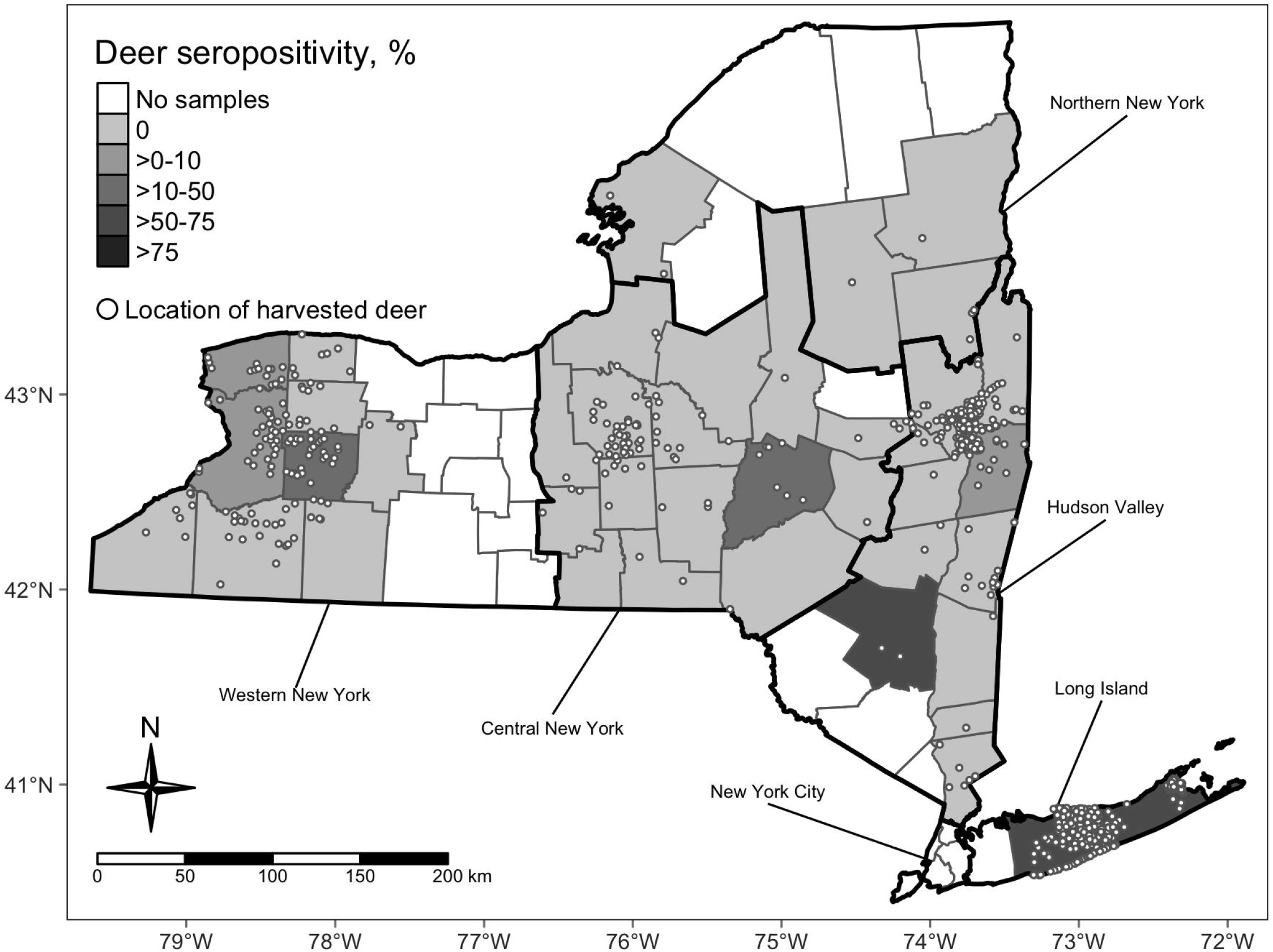
Map of New York State indicating sampling effort of hunter-harvested white-tailed deer bloods and BRBV seropositivity by county. Locations (open circles) of harvested deer are randomly jittered within townships.

**Figure 2.**
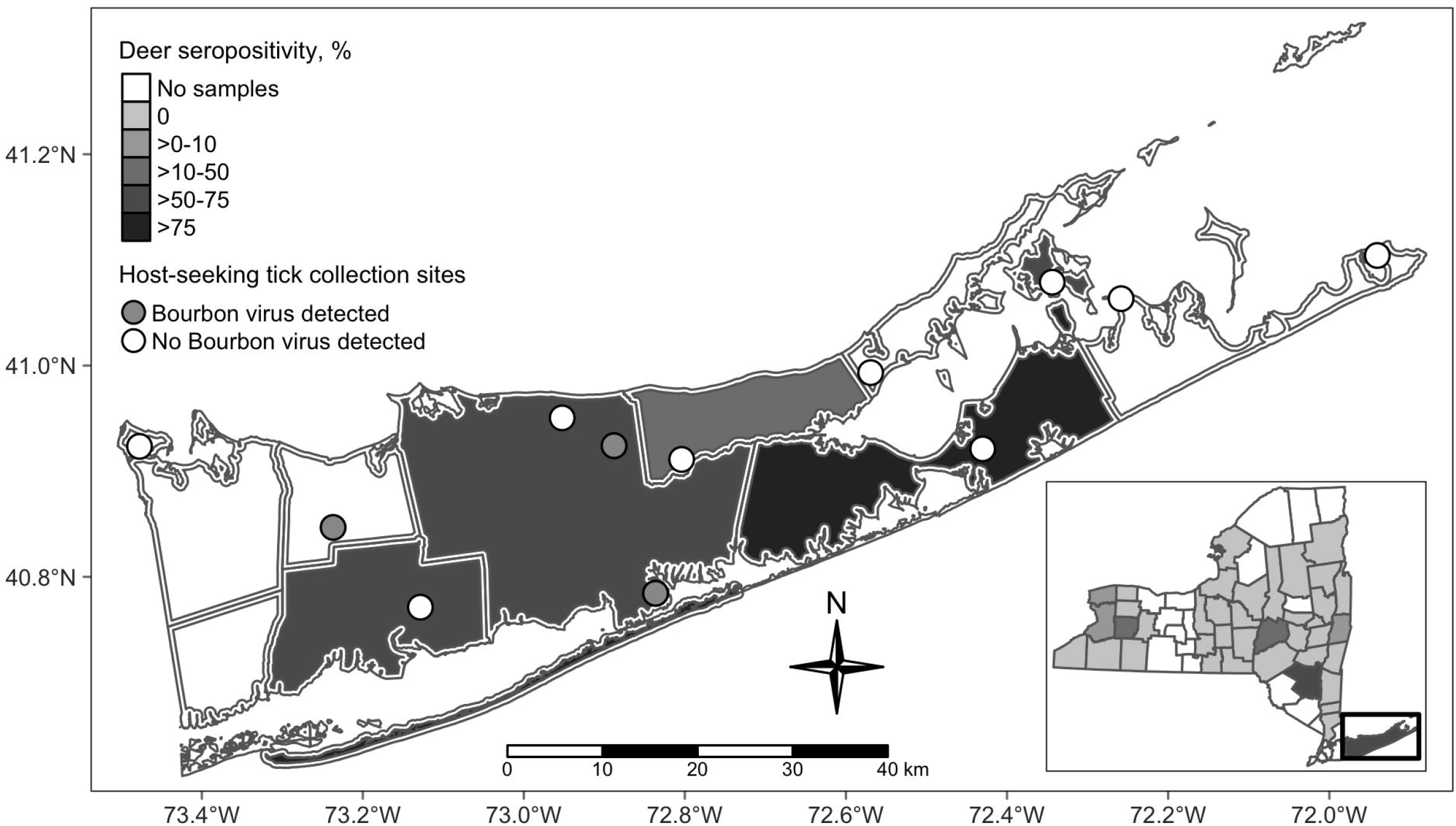
Map of Suffolk County, New York with inset of New York State. Circles within townships reveal tick collection sites. Open circles are sites with no evidence of BRBV. Dark circle represents approximate location of BRBV positive tick pools. BRBV seroprevalence is indicated by shading with the darker shades representing higher rates. Rates are depicted for Suffolk County townships, Fire Island or NY counties (map inset).

## Conclusion

Isolation of BRBV from the local tick population and high seropositivity in hunter-harvested white-tailed deer demonstrated active transmission throughout Suffolk County, NY since 2019. In addition, serologic evidence suggests the virus is present in other regions of NY. Consistent with previous BRBV field studies and the recent discovery of a closely related virus transmitted by *Amblyomma* species ticks in Japan, *A. americanum* ticks were implicated in local transmission of BRBV(2, 6). Tick minimal infection rates ranged from 0 to 0.77%. It is unclear whether the unengorged nymphs had acquired the virus as larvae feeding on viremic hosts, through co-feeding transmission or transovarially. These routes of transmission have been demonstrated in laboratory studies (5). BRBV was not detected in adult ticks tested during surveillance despite high numbers collected. Considering the overlap of adult and nymphal tick activity on Long Island, the effect of phenology on BRBV transmission will be the subject of further exploration during future surveillance campaigns.

Interestingly, of the five BRBV positive pools detected by the TickReport primer set, only one pool was positive with the primer set used during previous surveillance seasons despite similar assay limits of detection, suggesting genetic differences in the primer target regions. Phylogenetic analyses and *in vitro* growth characteristic studies are planned.

White-tailed deer are a sensitive sentinel model for many arboviruses given overall abundance and distribution, small home ranges, and the frequency on which they are fed upon by ticks and other hematophagous arthropods (14, 15). Suffolk County deer seropositivity (66.5%) was higher than the seroprevalence reported in North Carolina deer (56%), but lower than deer harvested in Missouri (86%) (8, 9). BRBV seroprevalence rates of white-tailed deer harvested from various areas in Suffolk County (Table 2) were similar to Oz virus seroprevalence rates (30.0-73.7%) in wild sika deer (*Cervus nippon*) sampled from Japanese prefectures located nearest to the initial detection of the virus (3). The lower seroprevalence in regions of NY outside of Long Island is likely attributed to fewer established populations of *A. americanum* or incidental transmission by bird-dispersed immatures originating from established regions. To date, no competent vertebrate host, including deer, has been implicated in BRBV amplification.

Results presented in this report emphasize the need to include emerging pathogens (BRBV, HRTV) in surveillance programs wherever lone star ticks are distributed. Clinicians outside of the Midwestern United States should be aware of the potential for human disease. It is unclear if the symptoms that the individual who removed the BRBV-positive tick experienced were the result of potential infection with BRBV, *Ehrlichia ewingii*, or an unrelated etiology, as patient blood samples were not available. Considering overlapping symptomologies (fever, fatigue, loss of appetite, thrombocytopenia and leukopenia) with other tick-borne infections including ehrlichiosis and Heartland virus disease, diagnoses can be challenging without specific testing. Currently, testing is only available at the Centers for Disease Control and Prevention and a few state health laboratories. Providers should request BRBV and HRTV testing for patients with history of tick exposure or travel to regions where *A. americanum* is reported and displaying clinical symptoms, including leukopenia and thrombocytopenia, that do not respond with antibiotic treatment.

## Data Availability

All data produced in the present work are contained in the manuscript

## Acknowledgments

The authors thank Steve Young, Josh Dwyer, Julia Goldstein, Sean Reagan, Emalee Clark, and Dylan Bartlett at SCDHS and, Lauren Rose, Anna Perry, and Jessica Stout at Wadsworth Center for assistance with tick surveillance, tick processing, and testing. The authors thank Beau Payne, Alexander Novarro, Jordan Raphael, and Kelsey Taylor for assistance with deer blood collections. Cells for serologic assays and media production were provided by the Wadsworth Center Media and Tissue Culture Core. This publication was supported by cooperative agreement U01CK000509, funded by the Centers for Disease Control and Prevention and by the National Institutes of Health (R01AI142572). Its contents are solely the responsibility of the authors and do not necessarily represent the official views of the Centers for Disease Control and Prevention, the Department of Health and Human Services or National Institutes of Health.

## Biographical sketch

Alan Dupuis is a Research Scientist at the Wadsworth Center, New York State Department of Health. His research interests include the role of the vertebrate host in the ecology of mosquito- and tick-borne viruses.

## Notes

### Competing Interest Statement

The authors have declared no competing interest.

### Funding Statement

This study was supported by cooperative agreement U01CK000509, funded by the Centers for Disease Control and Prevention and by the National Institutes of Health (R01AI142572).

